# Cerebrovascular risk factors impact brain phenotypes and cognitive function in healthy population

**DOI:** 10.1101/2022.03.29.22273047

**Authors:** Bingli Li, Yiran Wei, Kaijia Zhang, Carola-Bibiane Schönlieb, James Rudd, Chao Li

## Abstract

Cognitive decline is a major characteristic of ageing. Studies show that cardiovascular risk factors (CVR) are associated with cognitive declines and brain phenotypes, but the causality between CVR and cognitive function needs further understanding. In this study, we seek to investigate the causalities between CVR, brain phenotypes and cognitive function. We first generate a general factor (gCVR) representing common CVR and a score representing the polygenic risk (PRS). We then identify phenotypes of brain and cognitive functions associated with gCVR and PRS. Moreover, we conduct causal mediation analysis to evaluate the indirect effect of PRS through CVR, which infers the causality of gCVR on brain phenotypes and cognition. Further, we test the mediation effect of gCVR on the total effect of brain phenotypes on cognitive function. Finally, the causality between CVR and brain phenotypes is cross validated using Mendelian randomization (MR) with genetic instruments. The results show that CVR mediates the effect of PRS on brain phenotypes and cognitive function, and CVR also mediates the effect of brain phenotypes on cognitive changes. Additionally, we validate that the variation in a few brain phenotypes., e.g., volume of grey matter, are caused by CVR.

## Introduction

Cognitive health is of crucial significance with the increasing global burden of age-related diseases^1^. There is an urgent demand to improve our understanding of cognitive health, especially among the middle-aged and elderly population^2^. Mounting research has focused on understanding mild cognitive impairment (MCI) and dementia^3^, where many societal and physical factors are identified that contribute to cognitive health. As dementia still lacks effective treatments, and existing evidence suggests that a preclinical stage exists before the symptom onset, it is crucial to understand the risk factors leading to cognitive decline in a healthy population, which could help prevent the pathological cognitive decline and change disease trajectory^4^.

Enormous efforts have been made to identify modifiable risk factors, where cardiovascular risk (CVR) factors are one of the most recognized factors^4-10^. For example, a meta-analysis summarized the modifiable predictors of MCI and reported that diabetes could increase the risk of MCI^11^. Another meta-analysis indicated that cholesterol is a risk factor for cognitive decline^12^. On the other hand, studies have found that cardiovascular diseases may lead to brain structural alterations in populations with higher cardiovascular disease risk^13^. A recent study of 9,722 healthy individuals from the UK Biobank thoroughly examined the association between CVR factors and brain structure, which found higher levels of CVR factors associated with poorer brain health, indicated by global and regional grey and white matter macrostructure and microstructure^13^. Lastly, studies show that brain structure and cognitive function are closely related, and certain brain structures could significantly influence cognitive function^14^.

Existing studies, however, may not address the association and causal mechanisms among the CVR factors, brain structure and cognitive function for better understanding the mechanism of CVR affecting cognitive health. A study involving 94 non-demented/non-depressed older American adults found CVR factors (quantified by Framingham 10-year stroke risk, FSRP-10) were associated with tract-based brain structural connectivity and the changes in brain structural connectivity mediated the association between CVR factors and cognition^15^. The evidence in this study could potentially be strengthened with higher statistical power.

In this study, we hypothesized that a bi-directional casual effect exists between CVR and brain phenotypes. At the same time, CVR could mediate the effect of polygenic risk of blood pressure and brain phenotypes on cognitive function. Using a linear model, we first identified the phenotypes of brain and cognitive functions significantly associated with CVR. Another association was then performed to validate if those significant phenotypes are also associated with the polygenic risk scores of blood pressure (BP-CVR). In addition, causal mediation analysis was used to test whether CVR mediates the effect of BP-CVR on brain phenotypes and cognitive function. Further, we also tested the indirect effect of brain phenotypes through CVR on cognition using another mediation analysis. Finally, the casual effects between CVR/brain phenotypes and brain phenotypes/cognition were measured using Mendelian Randomization.

## Methods

### Participants

We included the UK Biobank participants. Exclusion criteria include self-reported or diagnosed neuropsychiatric disorders affecting cognition. The cohort was divided into imaging and non-imaging groups according to the availability of the image-derived phenotypes (IDPs) from the UK Biobank. Participants with self-reported neurological diseases are excluded. In summary, the imaging group includes 29,724 participants, and the non-imaging group consists of 227,273 participants. All imaging data were collected using the same model of MRI scanners. Behavioural and cognitive data were acquired using the same protocols by the UK Biobank. Written consent was obtained from all participants. Cognitive and neuroimaging data acquisition were conducted under standard protocols. This research has been conducted using data from UK Biobank, a major biomedical database www.ukbiobank.ac.uk. Written consent was acquired for all participants. Data acquisition and analyses in the present study were conducted under UK Biobank application 52802.

### Latent factors of cardiovascular risk factors and blood pressure

Confirmatory factor analysis (CFA) was conducted on cardiovascular risk (CVR) factors using the “lavaan” package^16^ in R to extract a latent variable for the cardiovascular risk (gCVR) for both imaging and non-imaging groups. The CVR factors include waist circumference, body mass index (BMI), diastolic blood pressure, systolic blood pressure, smoking status, diagnosis of hypertension, hypercholesterolaemia and diabetes. A separate latent factor (gBP) representing systolic and diastolic blood pressure is also calculated for generating polygenic risk scores of blood pressure (BP).

### Genome-wide association studies (GWAS)

GWAS summaries of the IDPs are downloaded from the Oxford Brain Imaging Genetics Server ^17^. The non-imaging cohort is selected to generate GWAS summary for gBP and gCVR) and general cognitive function represented by the Fluid intelligence score. Imputed genotype data (BGEN) was used. Basic quality control (QC) of single-nucleotide polymorphisms (SNP) were conducted by the UK Biobank according to the following criteria: Hardy-Weinberg disequilibrium (p<10e-5), low minor allele frequency (<0.005), low imputation accuracy (<0.1) and low call rate (<95%). For sample QC, the current study removed the participants with gender mismatched genetic data (according to data Field 31, 22001 and 22019), non-white British ancestry (according to Field 22006) and missingness > 0.1 (according to Field 22005). Participants without relatives were retained, and only one participant was randomly retained from each family (according to Field 22021 and Category 263). After QC, the QCed sample list and SNP list were generated. PLINK 2.0^18^ was applied to convert the BGEN file into PLINK files and for GWAS on the QCed sample and SNP lists.

### Polygenic risk score of blood pressure

The polygenic risk score (PRS) of blood pressure (BP-PRS) was calculated using PRSICE 2^19^. The GWAS summary of gBP of the non-imaging group is chosen as the base dataset, while the genotype data of the imaging group was used as the target dataset. Clumping was applied to remove the SNPs in Linkage Disequilibrium (LD) with each other. Eight p-value thresholds (PT, p < 0.0005, p < 0.001, p < 0.005, p < 0.01, p < 0.05, p < 0.1, p < 0.5 and p < 1) were applied to select the SNPs for generating PRS.

### Association analysis

The generalized linear model (GLM)^20^ was used to conduct association analysis between the gCVR and all IDPs (except QC category and bulk data) or cognitive scores. The p values of the association results were corrected for multiple comparisons using false discovery rate (FDR) correction. The Significance level of the corrected p values is set to 0.05.

### Phenome-wide association studies

The phenome-wide association studies (PheWAS) were conducted between the BP-PRS and the significant IDPs/cognitive scores from the above association analyses. The GLM function in R was used to perform PheWAS. The BP-PRS was set as independent variables, while the IDPs and cognitive scores were dependent variables. Each phenotype of IDPs and cognition was tested for the association with BP-PRS, with covariates controlled, including ten genetic principal components, age, sex, and genotype array. The p values were corrected using FDR. PheWAS is regarded as significant for a specific phenotype when there are a minimum of four significant p values (p<0.05) in the association tests of the eight BP-PRS with different p-value thresholds.

### Mediation analysis

Following the PheWAS, we perform mediation analysis to investigate whether the gCVR mediates the effect of BP-PRS on the IDPs and cognitive function using the structural equation model in the ‘lavaan’ package of R. The BP-PRS is defined as the predictor, and the gCVR is defined as the mediator. Only the significant phenotypes of IDPs and cognition from PheWAS analysis were defined as outcomes. Standard errors were bootstrapped 1000 times. The same covariates with the PheWAS were applied. The p values were corrected using FDR and the defined significance following the same rules of PheWAS with a minimum of four out of eight p values significant in the mediation analysis.

Another mediation analysis was applied to test whether the CVR mediates the effect of IDPs on cognitive function. Only the significant brain phenotypes and cognition scores from the PheWAS were defined as predictors and outcomes, respectively, while the gCVR was defined as the mediator. The same setup was applied as the mediation analysis of BP-PRS. The P values were corrected for multiple comparisons with FDR corrected.

### Mendelian randomization

Finally, Mendelian randomization (MR) was used to test the causal effects between CVR, IDPs and cognitive function. We used the “twosampleMR” package for conducting the two-sample MR analyses^21^. GWAS summaries of the gCVR from the non-imaging group and IDPs from the imaging group were used as the input for two-sample two-directional MR. Only the significant brain phenotypes of PheWAS were selected for testing the causal effects using the MR test.

To test the causal effect of CVR on IDPs, genetic instruments were selected from the GWAS summary of CVR at P < 5e-8. The corresponding SNPs in IDPs are extracted and harmonized to match the effect alleles. The clumping distance was set as 3000kb, and the LD threshold was set as 0.001. To test the causal effect of IDPson CVR, genetic instruments were selected from the GWAS summary of CVR at P < 8e-6. A less strict p-value threshold was applied to maintain the power of the MR test^22^. Other parameters were the same as the opposite direction MR test. MR Egger, IVM and weight median of MR methods were applied to test the significance of the two-sample MR.

## Results

### Participant characteristics

The characteristics of the participants in the imaging group and non-imaging group are shown in Table 1.

**Table 1.** Participants characteristics.

| Variable | Units | Descriptor<br>(imaging) | Descriptor<br>(non-imaging) |
| --- | --- | --- | --- |
| **Demographics** |  |  |  |
| Age | Years Mean (SD) | 55.08 (7.38) | 56.87 (8.03) |
| Sex | F(%F) | 10678 (51.22) | 121778 (53.60) |
| **CVRs** |  |  |  |
| Waist | cm Mean (SD) | 87.98 (12.55) | 90.12 (13.26) |
| BMI | Kg/m2 Median(IQR) | 25.87 (5.3294) | 26.67 (5.60) |
| BP_distolic | mmHg Mean (SD) | 78.55 (10.00) | 82.38 (10.05) |
| BP_systolic | mmHg Mean (SD) | 138.1 (18.38) | 138.6 (18.56) |
| SmokingPackPerYear | Packs Mean (SD) | 4.442 (11.08) | 6.571 (14.02) |
| Hypertension | Yes (Yes%) | 5544 (18.65) | 56841 (25.01) |
| Hypercholesterolaemia | Yes (Yes%) | 2631 (8.85) | 26873 (11.83) |
| Diabetes | Yes (Yes%) | 607 (2.04) | 7885 (3.47) |

### Cardiovascular risk is associated with IDPs and cognitive function

The association between the gCVR and IDPs are shown in Figure 1. In total, 843 out of 890 IDPs are significantly associated with the latent factors of CVR. The significantly associated IDPs include 656 diffusion MRI derived white matter phenotypes, 11 T2*-derived, 160 T1- derived regional volumes, 15 task functional MRI derived activation and one white matter hyperintensity volume. The white matter phenotypes contribute to the most significant proportion of brain phenotypes associated with the latent CVR, including 74 fractional anisotropy (FA), 69 intracellular volume fraction (ICVF), 73 isotropic volume fraction (ISOVF), 73 diffusion tensor mode (MO), 74 orientation dispersion (OD) and 293 mean diffusivity (MD). The highest positive association is found between the gCVR and the volume of grey matter in the Middle Temporal Gyrus, temporooccipital part (β =0.638, P<0.0001), while the highest negative association is found between the gCVR and Median T2star in the hippocampus (β = -0.403, P<0.0001).

**Figure 1.**
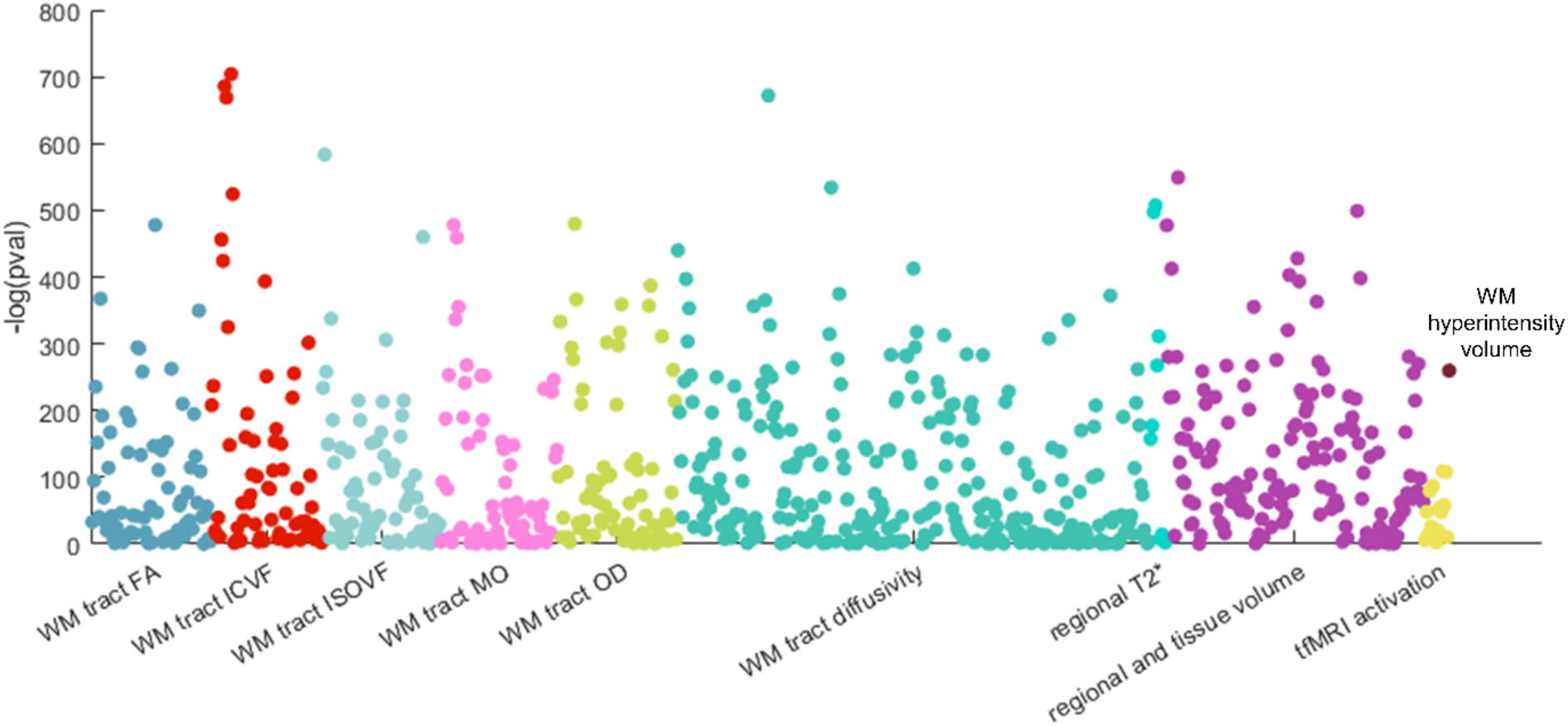
The significance plot for the association between CVR and IDPs. Dots in different colours indicate three different IDP categories. Each dot represents one phenotype. Only the IDPs surviving FDR corrected P-value < 0.05 are plotted.

In parallel, 33 of 84 cognitive functional scores are associated with the gCVR (Figure 2). The fluid intelligence contains the most significantly associated cognitive scores (5 scores). The symbol digit substitution has the highest negative association with gCVR (β = -0.482, P<0.0001) while the numerical memory (time number displayed) has the highest positive association (β = 0.248, P<0.0001). In general, the associations between cognitive function and CVR are less significant than those between CVR and IDPs.

**Figure 2.**
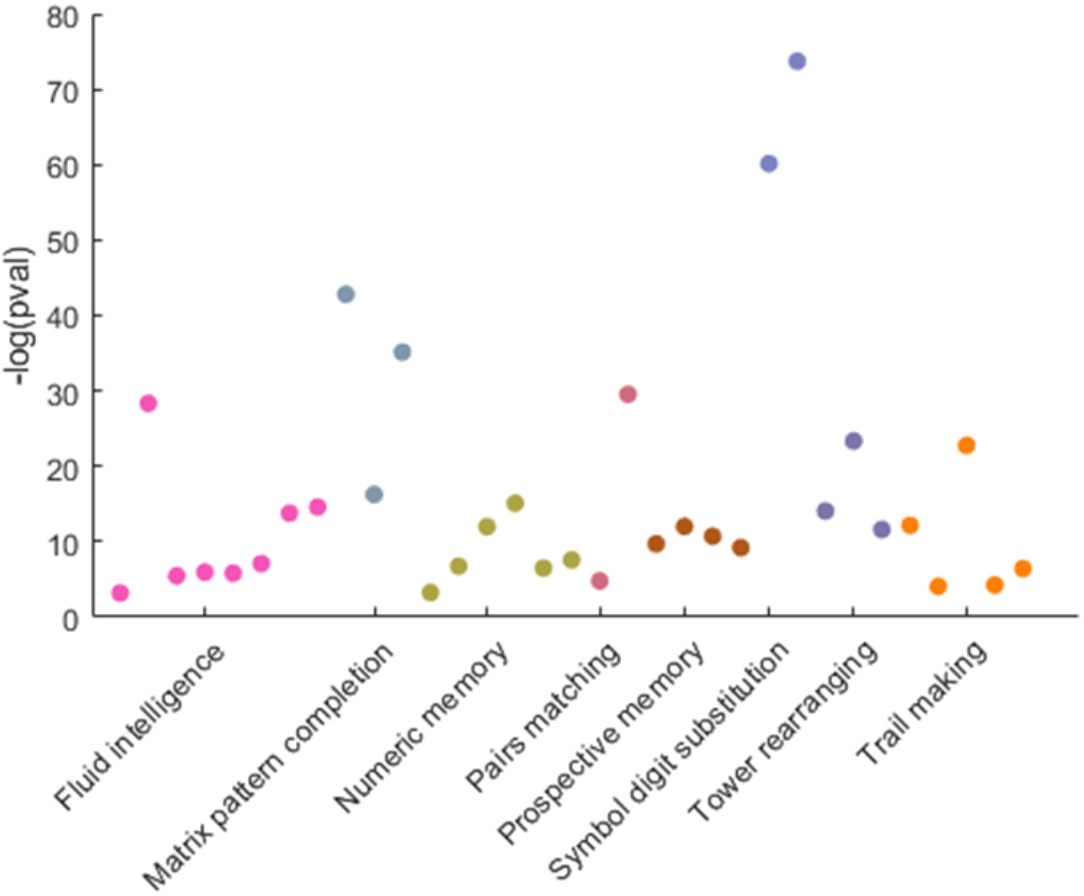
Significance plot for association between CVR and cognitive function scores. Dots in different colours indicate three different cognitive domains. Each dot represents one cognitive function test. Only the IDPs surviving FDR corrected P-value < 0.05 are plotted.

### Polygenic risk score of blood pressure is associated with IDPs and cognitive function

After selecting the CVR associated IDPs and cognitive scores, we performed PheWAS to investigate whether those phenotypes are associated with eight polygenic scores of the latent blood pressures (BP-PRS) calculated from different thresholds. We examine 876 phenotypes (843 IDPs and 33 cognitive functions). In total, 30 phenotypes (26 brain phenotypes and four cognitive functions) are shown significantly associated with BP-PRS at a minimum of four p-value thresholds after FDR correction for multiple comparisons.

Fig.3A-B present two examples indicating the p values of the association between the IDPs and BP-PRS of two PT (< 0.0005 and < 0.05). The overall significant plot (Fig. 3C) shows the association between IDPs and BP-PRS of all eight thresholds. In contrast to the dominant percentage (77.9%) of white matter phenotypes in the association between CVR and brain phenotypes, a higher percentage (61.5%) of grey matter volumes are found with significant association with BP-PRS. Specifically, only the median T2star in the right caudate is associated with all the eight BP-PRS. The volumes of grey matter in the right VIIIa cerebellum have the most significant associations (4 significant PT <0.001 and 2 significant PT < 0.05). For the white matter tract, the most significant IDPs are mean L1 in superior cerebellar peduncle (right) and mean L2/ISOVR in pontine crossing tract with five significantly associated BP-PRS.

**Figure 3.**
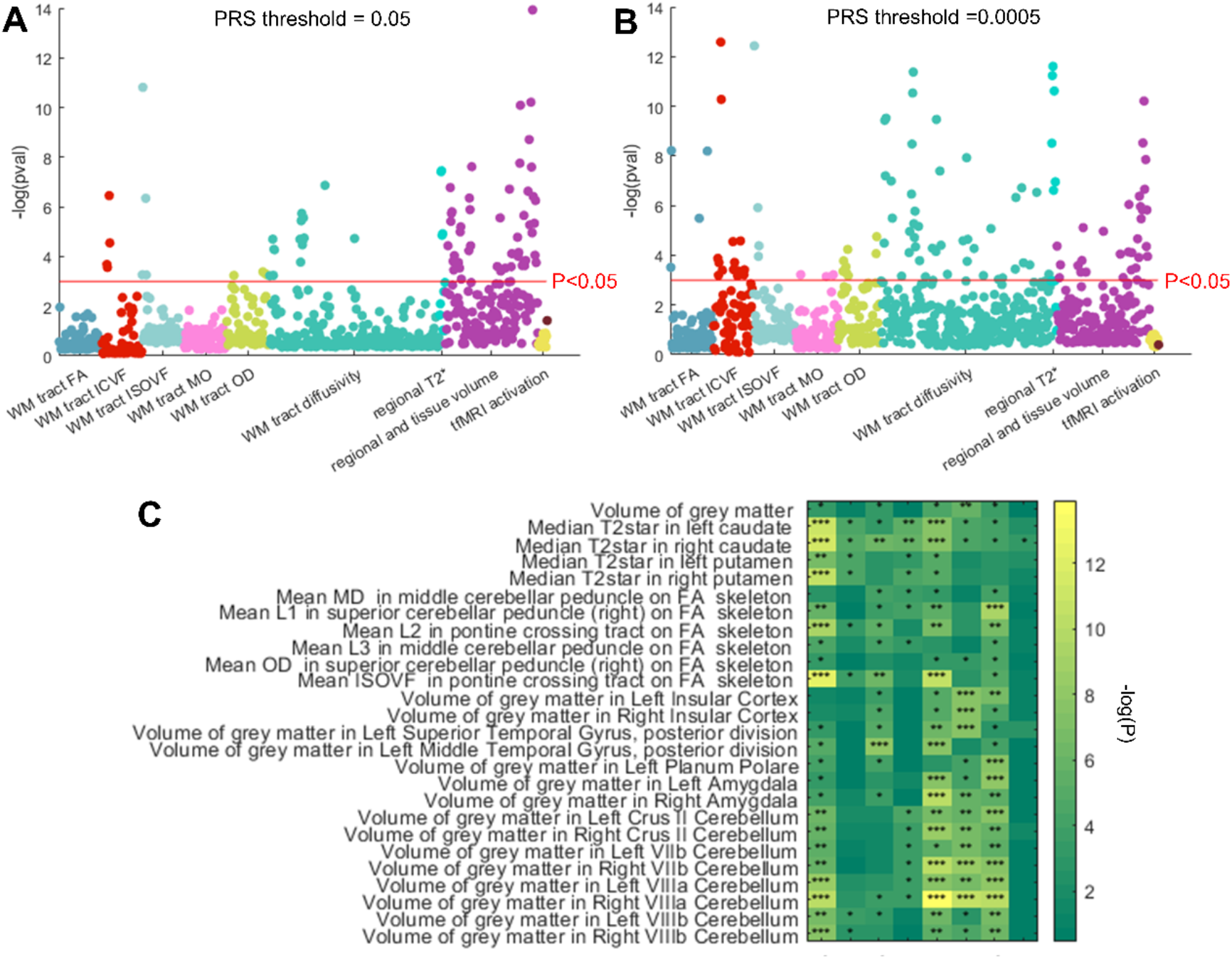
PheWAS between BP-PRS and IDPs. **A-B**: Significance plot for PheWAS results with PRS threshold of 0.05 and 0.0005. **The** significant IDPs from association analysis are selected for PheWAS with BP-PRS. The red line indicates the FDR corrected threshold of significance. **C**. Heatmap for the significant IDPs from the PheWAS results, significantly associated with BP-PRS. The colorbar indicates the negative log of p values. The asterisks indicate the significant level: ***P<0.001. **P<0.005, *P<0.05. The colormap indicates the -log(P).

For the associations between the gCVR and cognitive function, Fig.4A-B shows the examples of associations between cognitive function and BP-PRS of different thresholds. Fig. 4C shows the overall association between cognitive function and BP-PRS under eight thresholds. Pair matching (3 significant PT), numerical memory (5 significant PT), matrix pattern completion (5 significant PT), and fluid intelligence (4 significant PT) are the only cognitive scores associated with more than four BP-PRS. The most significant association is between fluid intelligence and BP-PRS at PT<0.01 with P = 2.02E-09.

**Figure 4.**
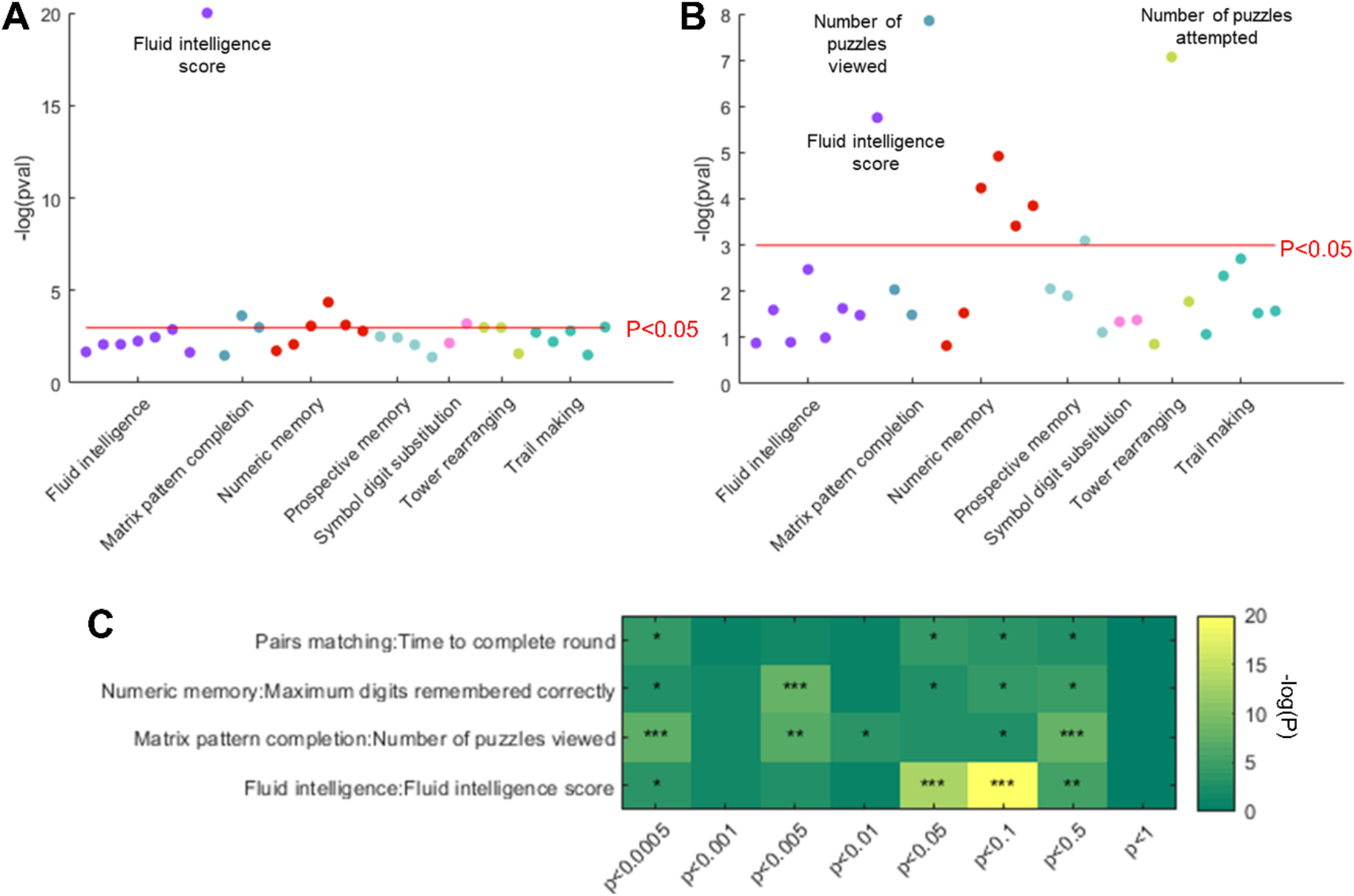
PheWAS between BP-PRS and brain phenotypes. **A-B**: Significance plot for the PheWAS results with PRS threshold at 0.05 and 0.0005. The significant cognitive function scores from association analysis are selected for PheWAS with BP-PRS. The red line indicates the FDR corrected threshold of significance. **C. Heatmap for the significant cognitive function scores from PheWAS results**. The shown cognitive scores are significantly associated with BP-PRS. The colormap indicates the negative log of p values. The asterisks indicate the significant level: ***P<0.001. **P<0.005, *P<0.05. Colormap indicates the -log(P).

### CVR factors mediate the effect of PRS of blood pressure on IDPs and cognitive function

To investigate whether the association between BP-PRS and IDPs and cognitive function are directly caused by the BP-PRS itself or mediated by the CVR. We performed the causal mediation analysis. The mediation results (Fig. 7) indicate that the CVR mediates all significant associations between the BP-PRS and IDPs. Specifically, positive indirect effects of BP-PRS through CVR are found on most brain phenotypes except for mean OD in the superior cerebellar peduncle. Strong mediation effects were found on phenotypes of both grey matter (Right VIIIa cerebellum) and white matter (Mean L2 in pontine crossing tract on FA skeleton). The BP-PRS with a PT of 0.0005 has the highest indirect effect via CVR on different IDPs. No significant indirect effects via CVR are found for BP-PRS with a PT of 1.

**Figure 5.**
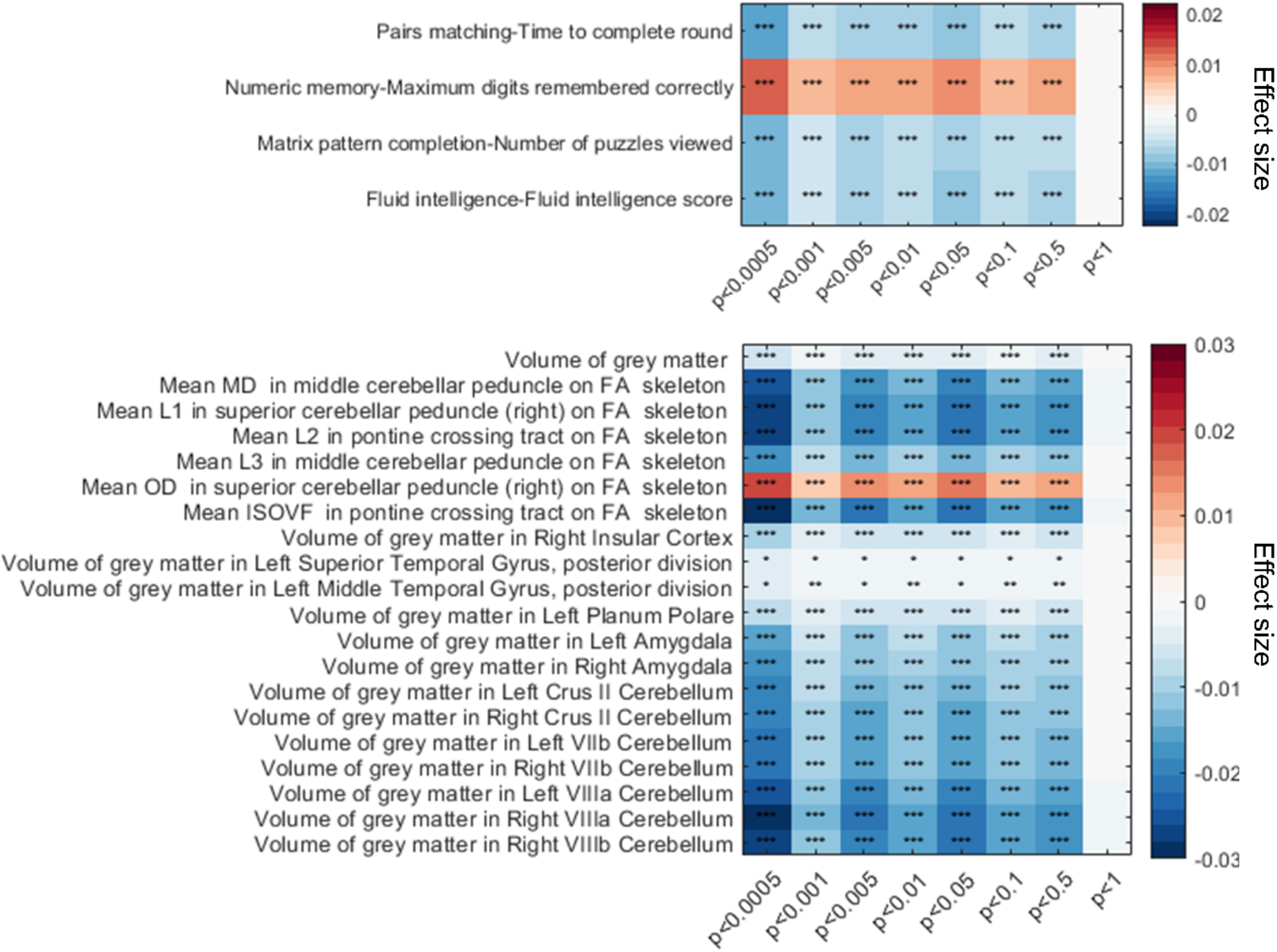
Indirect effect of BP-PRS through CVR on cognitive function (top) and IDPs (bottom). The asterisk indicates the significant level: ***P<0.001. **P<0.005, *P<0.05. The colormap indicates the effect size.

**Figure 6.**
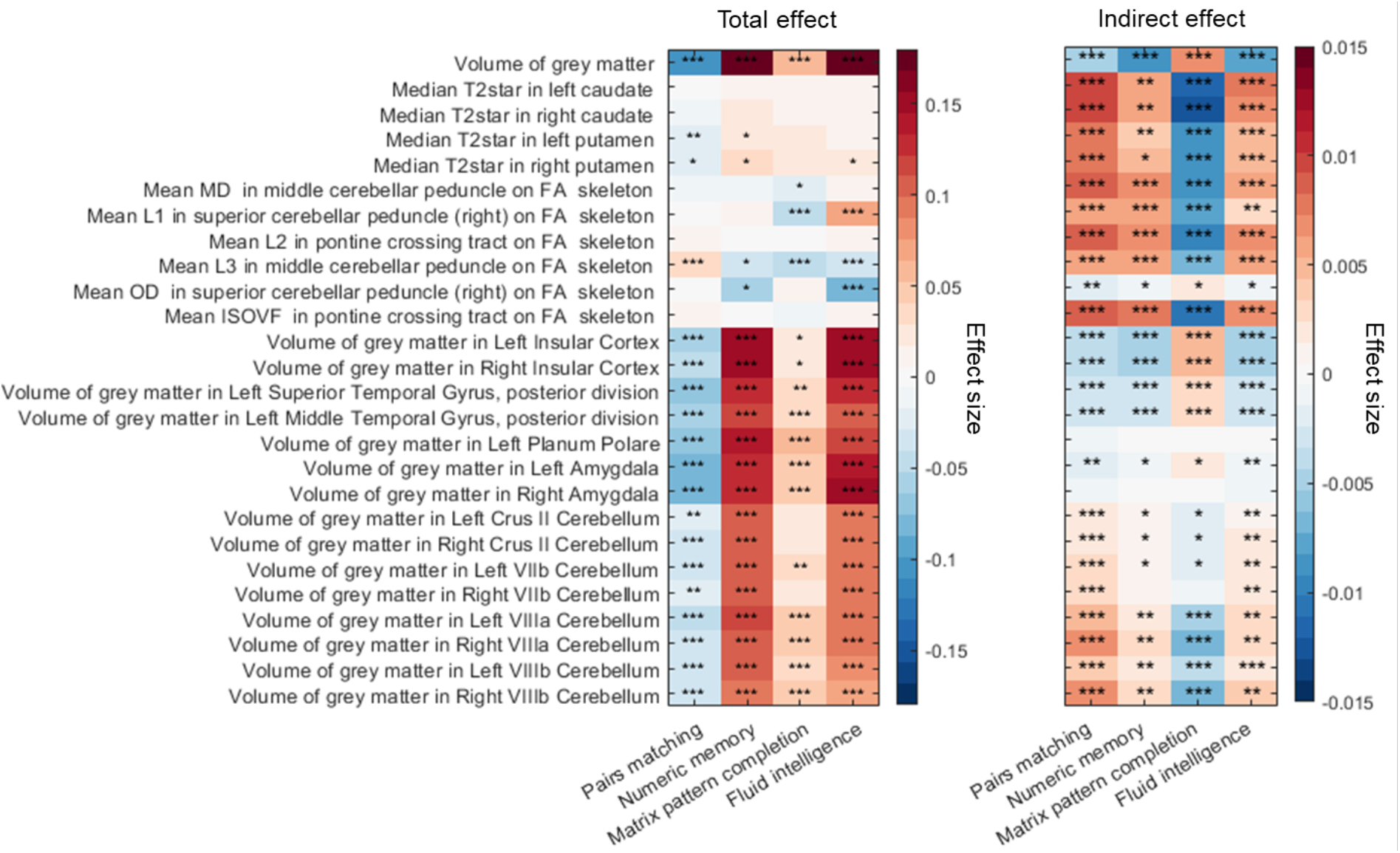
Total effects (left) and indirect effects (right) of brain phenotypes through CVR on cognitive function and IDPs. The asterisk indicates the significant level: ***P<0.001. **P<0.005, *P<0.05. The colormap indicates the effect size.

**Figure 7.**
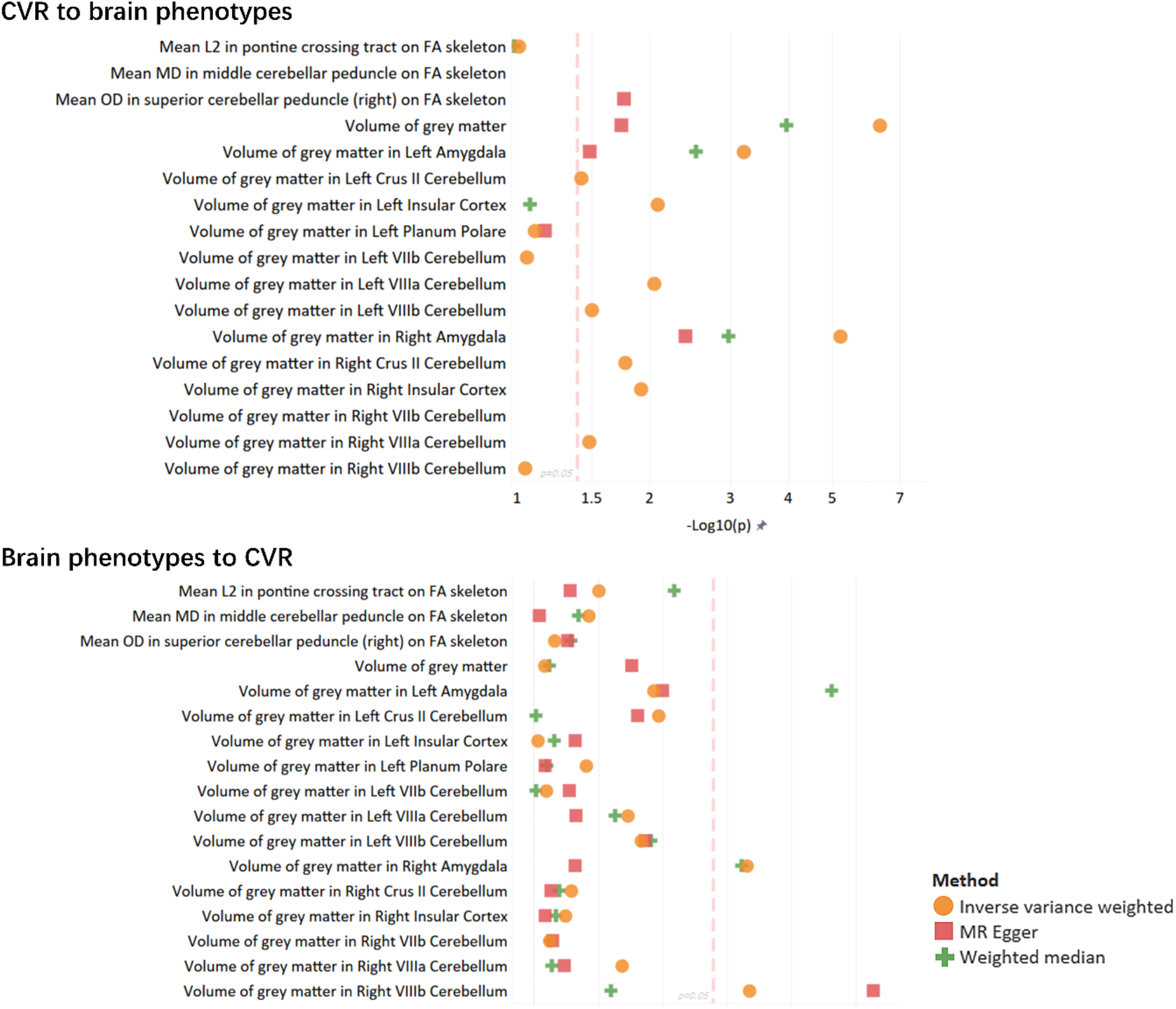
Bi-directional MR between CVR and brain phenotypes. Red lines indicate the significant threshold.

**Figure 8.**
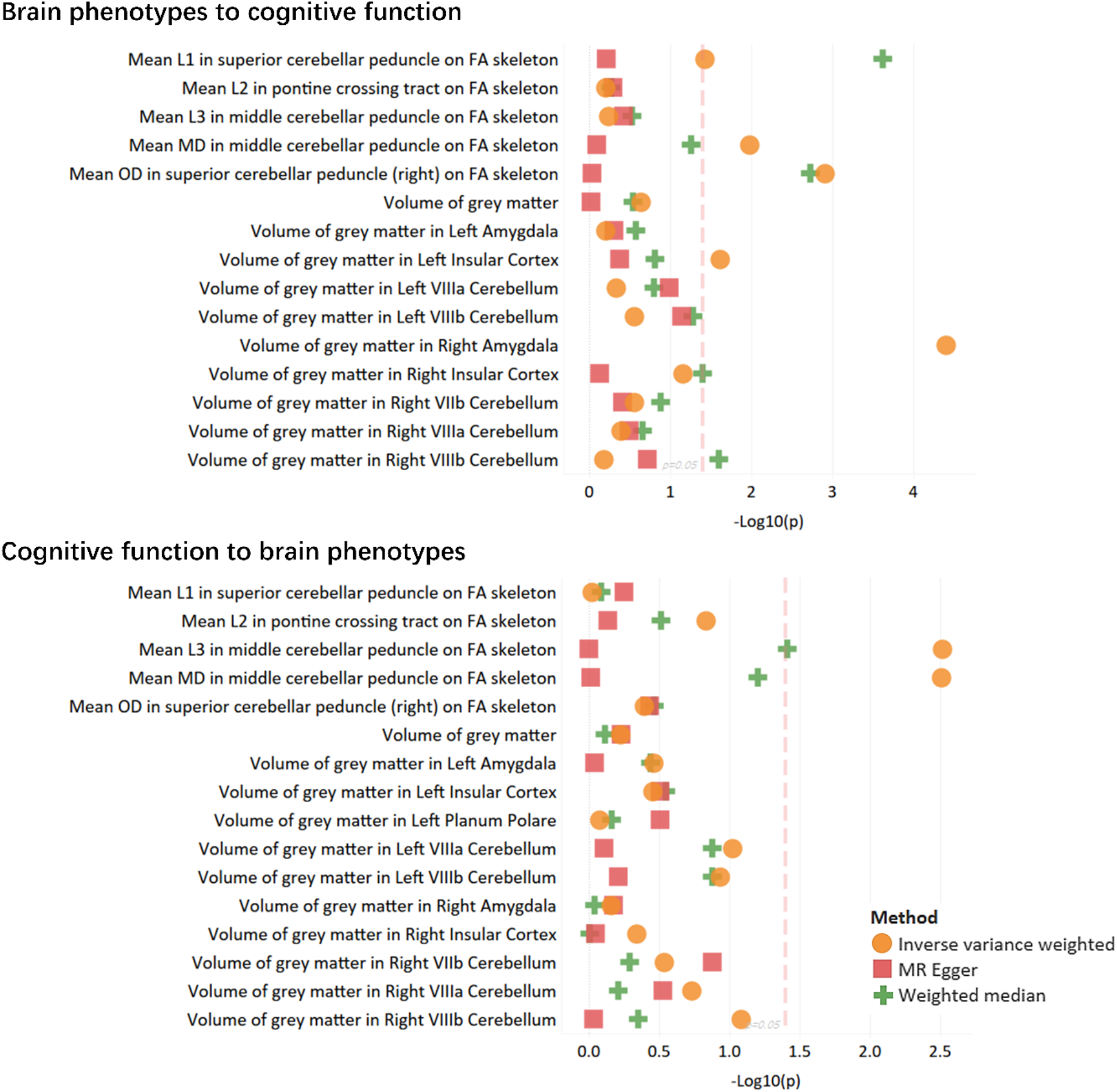
Bi-directional MR between brain phenotypes and fluid intelligence. Red lines indicate the significant threshold.

Similarly, significant mediation effects of CVR on cognitive function are also observed (Fig. 7). Negative mediation effects of CVR are found on pair matching, matrix pattern completion and fluid intelligence, whereas positive mediation effect is found on numerical memory.

### CVR factors mediate the effect of IDPs on cognitive function

To investigate whether the CVR also mediates the effect of IDPs on cognitive function, we performed another mediation analysis. The brain phenotypes and cognitive scores are chosen if significant in the PheWAS results. Our results show that the grey matter volumes have significant positive total effects on all cognitive function scores except pair matching. In contrast, fewer T2star and diffusion MRI derived phenotypes significantly affect cognitive function (Fig.6). In general, most IDPs have a negative total effect on pair matching and a positive total effect on numerical memory, matrix pattern completion and fluid intelligence. Specifically, the volume of grey matter has the highest positive total effect on maximum digits remembered correctly (numeral memory) (β = 0.176, P < 0.0001), fluid intelligence score (fluid intelligence) (β = 0.186, P < 0.0001) and number of puzzles viewed (matrix pattern completion) (β = 0.063, P < 0.0001), and the highest negative total effect on time to complete round (pairs matching) (β = -0.100, P < 0.0001).

For the indirect mediation effect of IDPs via CVR on cognitive function, all phenotypes derived from the T2star and diffusion MRI are found significant (P<0.001). The highest indirect effects include volume of grey matter on both numerical memory (β = -0.008, P < 0.0001) and fluid intelligence (β = -0.008, P < 0.0001), median T2star in right caudate on matrix pattern completion (β = -0.012, P < 0.0001). The Total effects generally have more significant effect size than indirect effects, whereas there are more significant indirect effects than significant total effects.

### Bi-directional two-sample MR

In addition to the causal mediation analysis, bi-directional two-sample MR tests were performed between CVR, IDPs and cognitive function. Only significant brain phenotypes from the PheWAS and fluid intelligence were chosen for this test.

A total of 13 of 27 brain phenotypes are found, with a minimum of one MR method showing a significant causal effect of CVR on IDPs. Three IDPs are found significant using all three MR methods, including volume of grey matter (β = 0.231 to 0.361, P = 4.73E-07 to 0.037), volume of grey matter in left amygdala (β = 0.174 to 0.324, P = 0.0006 to 0.032) and volume of grey matter in right amygdala (β = 0.200 to 0.381, P = 6.58E-06 to 0.019). The highest significance is found in inverse variance weighted methods (β = 0.258, P = 4.73E-07) for the volume of grey matter.

Only four phenotypes are found for the reverse effect of IDPs on CVR, with a minimum of one significant result. In comparison, only two phenotypes of the volume of grey matter in the right amygdala (β = -0.123 to 0.150, P = 0.022 to 0.024) and volume of grey matter in right vIIIb cerebellum (β = 0.060 to 0.161, P = 0.0008 to 0.020) are found with a minimum of two significant MR methods.

For the causal effect of IDPs on fluid intelligence, seven phenotypes are found with at least one significant result. In comparison, only three phenotypes of the same white matter (right superior cerebellar peduncle) are found significant in two MR methods (mean L1: β = 0.419 to 0.739, P = 0.0002 to 0.044, mean OD: β = 0.460 to 0.485, P = 1.2E-03 to 1.8E-03). As for the reverse effect of fluid intelligence score on brain phenotypes, only two phenotypes are found with at least one significant result, which is mean L3 in middle cerebellar peduncle on FA skeleton (β = 0.046 to 0.051, P =0.003 to 0.038) and Mean MD in middle cerebellar peduncle on FA skeleton (β = 0.049, P = 0.003).

## Discussion

In this study, we identified the imaging derived phenotypes and cognitive function by performing association studies with the cardiovascular risk factors. The results show that white matter tract phenotypes derived from the diffusion MRI are the most sensitive traits associated with the CVR factors. Moreover, the regional volumes are the second most traits related to the CVR factors.

After obtaining the CVR factors associated with IDPs and cognitive function scores, we performed the PheWAS for the polygenic risk score of blood pressure (BP-PRS) and selected phenotypes. In contrast to the association analysis, more phenotypes of grey matter volumes are found significant than phenotypes of white matter tracts, which suggests the high susceptibility of grey matter to high blood pressure. Surprisingly, many phenotypes of white matter tract diffusivity are associated with CVR factors but not significantly associated with BP-PRS.

To investigate whether the variation of brain phenotypes and cognitive function is caused by the genetic risk of blood pressure or gCVR, we conducted the causal mediation analysis with BP-PRS as the predictor, gCVR as mediator and brain phenotypes/cognitive function scores as the outcomes. The significant indirect effects of BP-PRS via gCVR suggest the casual mediation effect on IDPs and cognitive function. Overall, the results show that the genetic risk of blood pressure does not necessarily cause a direct effect on brain phenotypes and cognition. In addition, we also found that gCVR mediates the total effect of brain phenotypes on cognitive function by applying another causal mediation analysis. Additionally, the weak total effect of white matter phenotypes suggests the weak association between white matter tracts and cognitive function. Finally, we tested the causal effect between brain phenotypes and CVR using the two-directional two-sample MR. The result shows a causal bi-direction impact between CVR and different brain phenotypes. This study validates our hypothesis that CVR plays an important role in the changes in brain phenotypes and cognitive function. The mediation effects of CVR significantly affect different cognitive domains.

Our study has limitations. Firstly, the CVR factors contain binary variables, e.g., hypertension, diabetes, hypercholesterolaemia, smoking packs per year, which may be simplified. Continuous characterization of CVR could be included in future studies to improve the CVR scores. Secondly, the GWAS summaries of IDPs are from a larger subgroup of UK biobank than our imaging group. Finally, lifestyle phenotypes, e.g., sleep, alcohol consumption, physical activity, associated with brain phenotypes and cognitive scores in previous studies, are not considered in this study. Future work may investigate the effect and interactions between CVR factors and lifestyle variables with a more complex structural equation model.

## Conclusion

The latent factors of aggregated cardiovascular risk factors have a significant association with IDPs and cognition, while significant bi-directional causalities are found between the IDPs and CVR factors. In addition, strong mediation effects of CVR factors are found in the indirect effect of blood pressure genetic risk score on IDPs and functions. Moreover, CVR factors are also found mediating the effect of IDPs on cognitive function.

## Data Availability

All data produced in the present study are available upon reasonable request to the authors.

